# Type-2 diabetes biomarker discovery and risk assessment through saliva DNA methylome

**DOI:** 10.1101/2024.12.20.24319403

**Authors:** Wenbin Guo, Marco Morselli, Kimberly C. Paul, Michael Thompson, Beate Ritz, Matteo Pellegrini

**Affiliations:** Bioinformatics Interdepartmental Program, University of California Los Angeles, Los Angeles, CA, 90095, USA; Department of Molecular, Cell, and Developmental Biology, University of California Los Angeles, Los Angeles, CA, 90095, USA; UCLA-DOE Institute for Genomics and Proteomics, University of California Los Angeles, Los Angeles, CA, 90095, USA; Department of Epidemiology, Fielding School of Public Health, University of California Los Angeles, Los Angeles, CA, 90095, USA; Department of Neurology, David Geffen School of Medicine, University of California Los Angeles, Los Angeles, CA, 90095, USA; Department of Environmental Health, Fielding School of Public Health, University of California Los Angeles, Los Angeles, CA 90095, USA

## Abstract

The rising prevalence of type 2 diabetes (T2D) motivates innovative strategies to deepen disease understanding and enhance diagnostic capabilities. This study measures diabetes-specific epigenetic signals in saliva, establishing saliva DNA methylome as a promising medium for T2D screening and study. By integrating comprehensive whole-genome bisulfite sequencing (WGBS) and high-depth targeted bisulfite sequencing (TBS), we developed a cost-efficient two-step approach to profiling DNA methylation at regions of interest. WGBS analysis confirmed T2D-specific methylation signatures in saliva, revealing their enrichment in immune and metabolic regulation pathways. TBS enabled accurate cell type deconvolution, revealing minimal differences in cellular composition between diabetic and non-diabetic samples, suggesting intrinsic molecular changes drive the observed methylation changes. Epigenome-wide association studies further identified significant CpG sites, notably in the *ABCG1* region, with strong potential for T2D status prediction. These findings validate the saliva DNA methylome as a scalable, non-invasive resource for T2D biomarker discovery, advancing opportunities in T2D screening, risk assessment, and personalized medicine.

## Introduction

Diabetes mellitus, a multifaceted metabolic disorder characterized by hyperglycemia, continues to pose a considerable and escalating global health challenge. According to the World Health Organization and the Centers for Disease Control and Prevention, the prevalence of diabetes has surged more than fourfold since 1980 [1], affecting approximately 529 million individuals worldwide [2] and 38.4 million in the United States in 2021 [3, 4]. Notably, over 90% of these cases are type 2 diabetes (T2D) [5]. This alarming rise (Figure S1) underscores the urgent need for deeper disease mechanism understanding, innovative diagnostic tools, as well as effective management strategies. Timely detection and intervention are crucial for managing diabetes, preventing associated complications, and reducing the economic burden on patients and healthcare systems.

Recent years have witnessed burgeoning interest in the role of epigenetics underlying diabetes [6, 7, 8], focusing on how environmental factors and lifestyle choices can induce gene expression changes without altering the DNA sequence. Among various epigenetic modifications, DNA methylation has garnered substantial attention for its robust and dynamic nature, playing important roles in gene regulation, cell differentiation, development and maintenance of homeostasis [9, 10]. Alterations in DNA methylation can contribute to disease and are often reflective of disease states, making them informative for disease mechanism research and diagnostic purposes [6, 11]. In the context of T2D, DNA methylation has been implicated in its onset [12, 13], progression [14], and complications [15, 16, 17], with emerging evidence highlighting its utility for diabetes risk prediction [18, 19]. Aberrant methylation patterns are also found in key genes associated with glucose metabolism [20], insulin secretion [21], insulin resistance [22], and inflammatory responses [23, 24]. These findings establish DNA methylation changes as valuable biomarkers for T2D, emphasizing their potential in elucidating disease mechanisms and developing novel diagnostic and treatment strategies.

Despite advancements in understanding DNA methylation changes in diabetes, most studies have focused on tissues such as blood, skeletal muscle, adipose tissue, and pancreas [6, 7, 8, 25, 26, 27, 28, 29], while the potential of saliva DNA methylation as a non-invasive biomarker remains underexplored. Saliva offers a particularly appealing option due to its ease of collection and high patient compliance, making it ideal for disease screening and routine monitoring. Recent studies have demonstrated a high similarity in methylation profiles between blood and saliva [30, 31], suggesting that disease-associated epigenetic signals identified in blood may also be detectable in saliva. This evidence forms the basis of our hypothesis that the saliva methylome can serve as a valuable medium for identifying T2D biomarkers. If validated, the saliva methylome profiles could facilitate T2D screening and monitoring, paving the road for future applications in T2D diagnostics and management.

A major challenge in current methylation profiling is the substantial resource demand, particularly with whole-genome bisulfite sequencing (WGBS), which remains prohibitively expensive for large-scale studies and clinical applications. While methylation microarrays offer a more affordable alternative and are widely used in DNA methylation research [32, 33], they capture only a limited, predetermined subset of CpG sites, potentially overlooking critical regions relevant to the disease of interest. Recognizing that many CpG sites exhibit minimal variation across cell types [34, 35] and non-cancer diseases [36], we identified an opportunity to reduce costs by selectively measuring the informative regions. In this study, we devised and implemented a cost-effective two-step strategy for T2D biomarker research (Figure 1). First, pooled WGBS of saliva DNA was conducted to identify T2D-associated signals, revealing 1,358 differentially methylated regions (DMRs) between diabetic and non-diabetic groups. Building on these findings, we designed custom probes to enrich these DMRs and other informative regions for targeted bisulfite sequencing (TBS). This integrated approach synergizes the broad genomic coverage of WGBS with the high-depth profiling of TBS, enabling precise DNA methylation measurements in genomic regions OF interest. By focusing sequencing efforts on relevant targets, this approach achieves cost-efficiency and makes large-scale study and routine screening more economically feasible.

**Figure 1:**
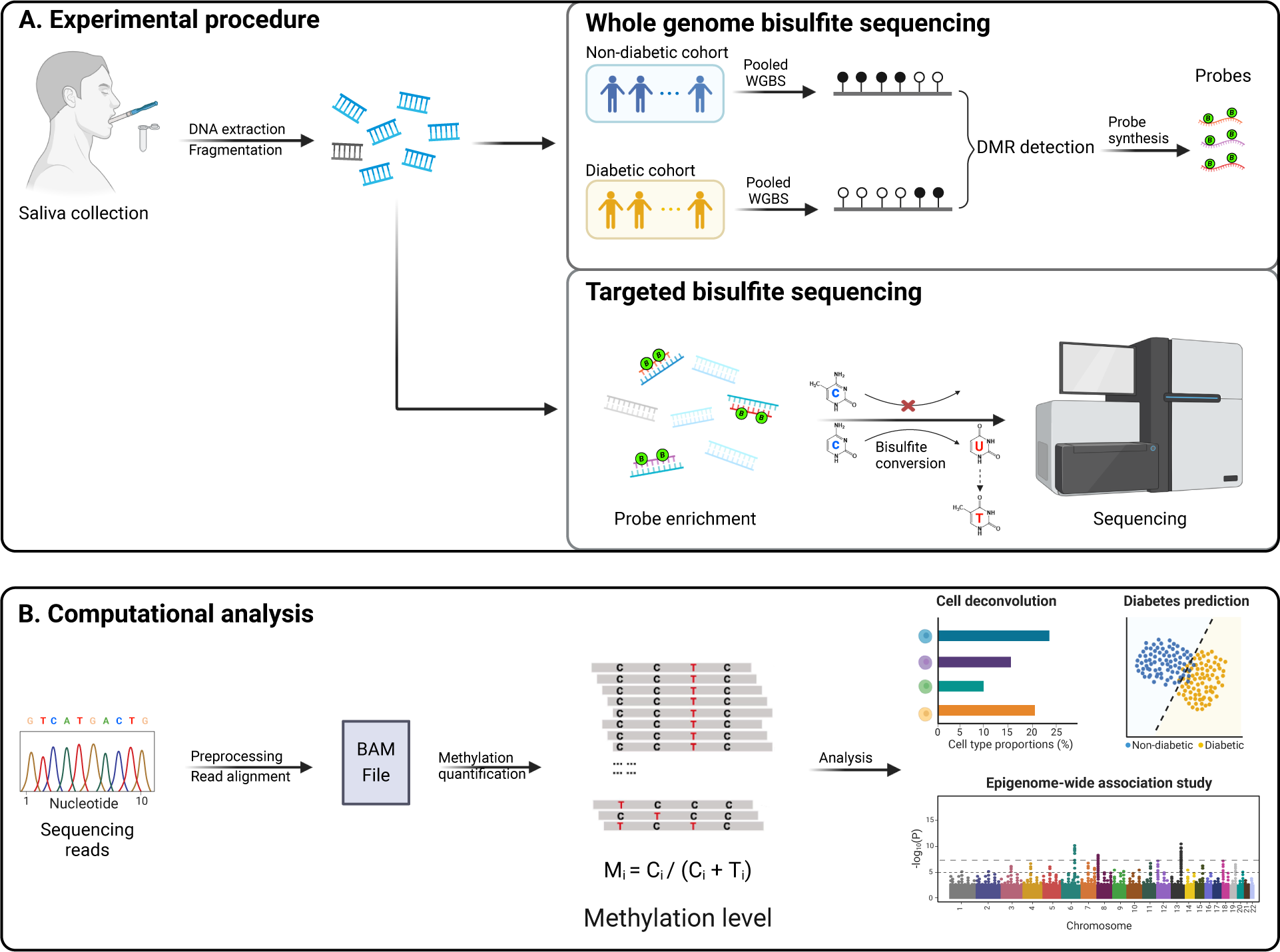
Study design for saliva DNA methylome analysis in Type 2 diabetes. (A) Experimental procedure. Participants’ saliva samples were collected, followed by DNA extraction and fragmentation. Pooled samples from non-diabetic and diabetic cohorts were then subjected to whole-genome bisulfite sequencing (WGBS) to identify differentially methylated regions (DMRs) associated with T2D. Probes targeting these DMRs were synthesized and used for targeted region enrichment, followed by bisulfite conversion and sequencing in high-efficiency Targeted Bisulfite Sequencing (TBS). (B) Computational Analysis. Sequencing reads underwent preprocessing and alignment, with methylation levels quantified as the ratio of methylated cytosine (C) counts to the total counts at each CpG site. The methylation data were used for downstream analysis, including cell type deconvolution, an epigenome-wide association study, and diabetes status prediction.

Our study validated the presence of T2D-associated signals in the saliva methylome for the first time and provided key biological insights into the molecular basis of T2D. WGBS analysis revealed that the identified DMRs were significantly enriched in immune and metabolic pathways, consistent with the established pathophysiology of T2D [37]. TBS provided a high-depth profiling of the targeted regions and allowed for accurate cell-type deconvolution. This analysis revealed no major differences in cell type composition between diabetic and non-diabetic samples, suggesting the observed methylation changes are likely driven by intrinsic molecular alterations rather than shifts in cellular proportions. To further investigate the molecular changes underlying T2D, an epigenome-wide association study (EWAS) was conducted on TBS data and identified 12 significant CpG sites with the top hit in the *ABCG1* region, replicating and reinforcing findings from previous blood-based studies [32, 38]. Collectively, these findings establish saliva as a robust and practical medium for T2D research, enabling the precise identification of T2D-associated biomarkers. By integrating WGBS and TBS, this approach provides a cost-efficient and scalable framework for large-scale screening and monitoring. This study underscores the transformative potential of saliva-based epigenetic approaches in advancing T2D research and diagnostic applications.

## Results

### WGBS identifies DMRs associated with diabetes in saliva

To investigate DNA methylation changes associated with T2D while optimizing sequencing efficiency, we implemented a carefully designed sample pooling strategy followed by Whole Genome Bisulfite Sequencing (WGBS). In this study, we pooled 96 saliva samples with matched demographical attributes into four groups: Diabetic Male, Diabetic Female, Non-diabetic Male, and Non-diabetic Female, with a balanced sample size per group. This pooling approach ensured adequate representation of each group and enabled robust comparisons across groups at a reduced cost. The WGBS data were then processed and aligned to the human reference genome (hg38) with CpG methylation levels quantified. Downstream differential methylation region (DMR) analysis between diabetic and non-diabetic groups revealed 1358 potential DMRs out of 162833 total regions (0.8%), visualized using a volcano plot (Figure 2A) and a heatmap (Figure 2B). These findings highlight significant epigenetic variations (both hypo- and hyper-methylation) between diabetic and non-diabetic individuals.

**Figure 2:**
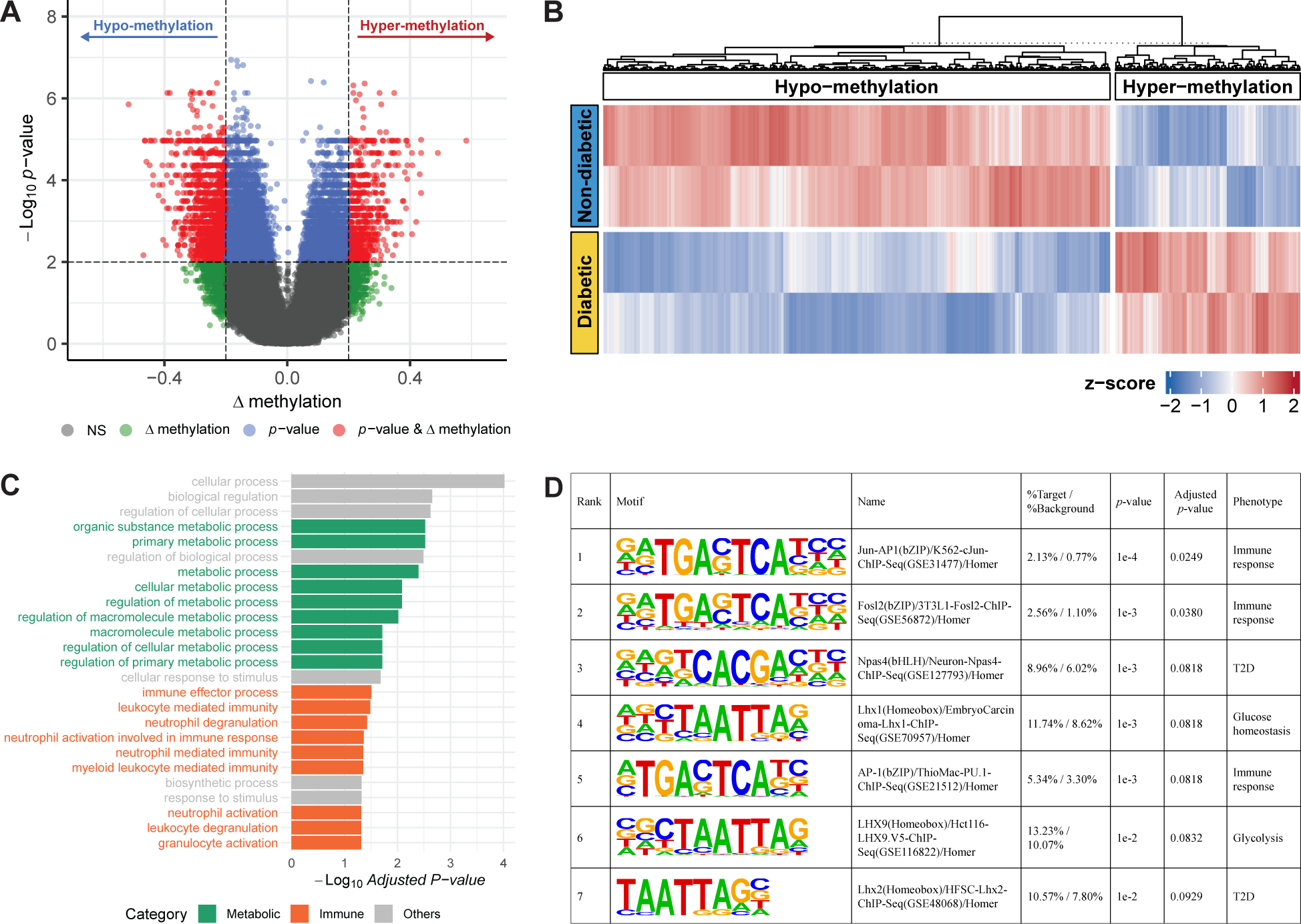
Differential methylation region and genomic region enrichment analysis for saliva WGBS data. (A) Volcano plot showing differential methylation region (DMR) analysis results, comparing diabetic group to non-diabetic controls. The x-axis represents the difference in methylation levels (Δmethylation), while the y-axis displays the −log10 p-values. Regions where both Δmethylation and the p-value exceed their respective thresholds are highlighted in red, representing hypo-methylation (left) and hyper-methylation (right). Regions where only the Δmethylation or p-value passe their corresponding threshold are shown in green and blue, respectively. Non-significant regions are depicted in gray. (B) Hierarchical clustering heatmap of DMRs’ methylation levels across diabetic and non-diabetic groups. The color scale represents z-scores, with hypo-methylated regions indicated in blue and hyper-methylated regions in red, highlighting differential methylation between the two groups. (C) Bar plot showing the genomic region enrichment analysis results of DMRs. The x-axis represents the −log10 adjusted p-value of enrichment, and the y-axis lists the enriched Gene Ontology (GO) terms of biological processes. Metabolic-related processes are highlighted in green, immune-related processes in orange, and others in gray, with notable enrichment in pathways related to cellular metabolic and immune responses. (D) Table summarizing the significantly enriched transcription factor binding sites. Each motif was ranked by significance, and the percentage of target versus background regions, p-value, adjusted p-value, and associated phenotype were provided.

Genomic region enrichment analysis was conducted to elucidate the biological relevance of the identified DMRs. The results exhibited substantial enrichment in genomic regions associated with metabolic regulation and immune response pathways (Figure 2C, Figure S2), underscoring their potential relevance in diabetes pathogenesis. Notably, several key pathways, such as leukocyte-mediated immunity and neutrophil activation, were significantly enriched, aligning with the current understanding of diabetes as a multifactorial disease involving intricate interactions between metabolic dysfunction and immune responses [37]. Based on the identified DMRs, we designed a set of probes (n=937) for targeted bisulfite sequencing. Motif analysis of these probe-enriched regions revealed seven significant transcription factor binding sites (p-value *<* 0.01, adjusted p-value *<* 0.1, Figure 2D), which are associated with T2D [39] and related traits, such as glycolysis [40] and immune response [41]. This association underscores the functional relevance of the identified DMRs and enriched regions with diabetes pathophysiology. Taken together, the WGBS analysis confirmed the presence of diabetes-specific methylation signals in saliva and facilitated the screening of genomic regions enriching these signals, paving the way for efficient profiling through Targeted Bisulfite Sequencing.

### TBS enriches target regions with high sequencing depth

To enhance efficiency in large-scale epigenetic profiling, we implemented targeted bisulfite sequencing (TBS) using a curated set of probes. This set includes probes designed to enrich the identified DMRs from WGBS analysis, as well as additional probes targeting regions associated with phenotypes such as aging, cell type, BMI and metabolic disorders [29, 42, 43]. A total of 8154 probes were used throughout the targeted bisulfite sequencing study, capturing ∼1M bases of the genome. Genomic coordinate overlap analysis with the existing EWAS database [44, 45] revealed that more than 40% of the probes overlap with known EWAS sites associated with diabetes and related traits, including BMI, obesity, fasting glucose levels, insulin levels and resistance (Figure 3A), ensuring the capture of the diabetes-informative methylation regions.

**Figure 3:**
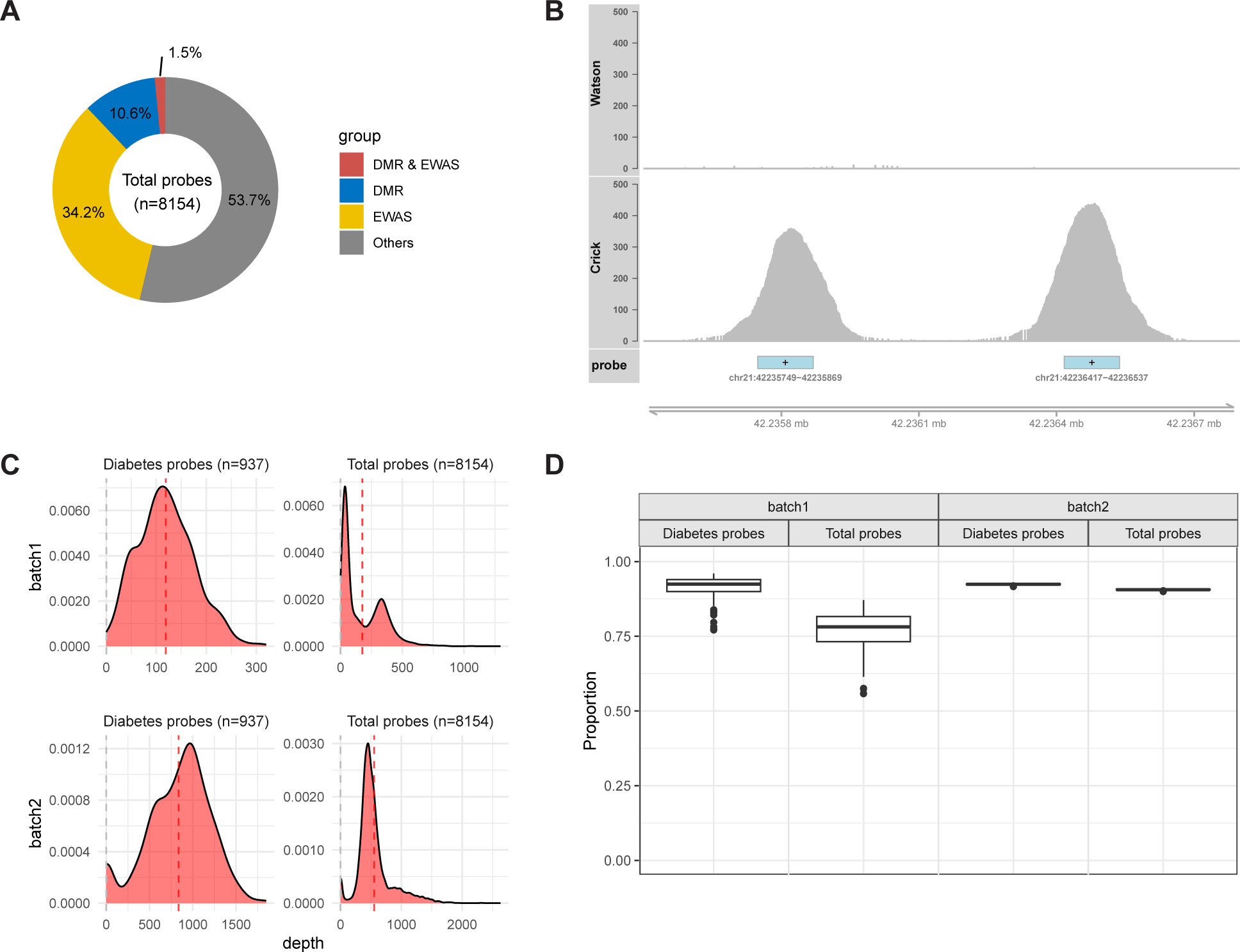
TBS captures desired region with high depth with reduced cost. (A) Pie chart illustrating the composition of the probe set (n=8154), highlighting its overlap with the differentially methylated regions (DMRs) identified in WGBS data and the public EWAS database. The probes are categorized as overlapping with DMR & EWAS (blue), DMR only (red), EWAS only (yellow), and other regions (gray). (B) Coverage plot showcasing an example of read coverage across a targeted genomic region (chr21:42,235,500-42,236,800) in a sample’s TBS data. The x-axis represents the genomic coordinates, and the y-axis shows the depth at each locus. Both Watson and Crick strands are displayed, with the targeted probe region highlighted in blue. The plus signs indicate probes designed on the Watson strand to capture the Crick strand. (C) Density plots showing the depth distribution of probes targeting diabetes DMR regions (n=937) and the total probe set (n=8154) across two batch samples. The red dashed line indicates the average depth of the enriched regions, with grey dashed lines indicating the non-enriched background regions. (D) Box plots displaying the percentage of CpG sites within the probe regions that achieve a sequencing depth greater than 10x. The plots demonstrate the efficiency of TBS in achieving high sequencing depth for the targeted regions across probe sets and batches.

With the curated probe set, we conducted targeted bisulfite sequencing on two cohorts (Supplementary Data 1), aiming for 10 million reads per sample. Our results confirmed that TBS can effectively capture the targeted genomic regions with high depth (Figure 3B). Of note, the enriched regions exhibited an average of 1300-fold higher depth than non-enriched background regions (Figure 3C, Figure S3), and over 80% of the CpG sites within the targeted regions had depth greater than 10 counts (Figure 3D). These findings demonstrate the remarkable efficiency of TBS in profiling targeted genomic regions with high depth while achieving cost efficiency. The successful capture of informative regions establishes TBS as a scalable solution for high-throughput epigenetic studies. Its high-depth coverage of CpG sites within targeted regions enables accurate and reliable DNA methylation quantification, ensuring robust statistical power for detecting differential signals in downstream analyses.

### Cell type deconvolution reveals minimal T2D-related compositional changes in saliva

Both the WGBS and TBS technologies are applied to bulk saliva samples, which obscures the specific cell type abundance associated with T2D in saliva. To address this, we first assessed whether the TBS sites contain cell type information. We downloaded a WGBS dataset containing a comprehensive methylation atlas of normal human cell types [46] and identified cell type-specific regions. By overlapping with the TBS sites, we found a significant proportion of the TBS sites fell within these cell type-specific regions, sufficiently distinguishing the different cell types in saliva tissue (Figure S4). To further validate the utility of these sites for cell type deconvolution, we generated in-silico mixtures of DNA methylation profiles with known cell type proportions. Using these simulated datasets, we performed cell type deconvolution analysis using the Houseman method [47] (Figure S5A), achieving a root mean square error (RMSE) of less than 0.01 and an R-squared value approaching to 1 (Figure S5B). Repeated experiments consistently showed high accuracy (Figure S5C), confirming that the TBS sites support accurate cell-type deconvolution.

Following this validation, we applied the deconvolution method to bulk saliva TBS data to investigate cell type composition in our samples. The analysis revealed that monocytes, granulocytes, and epithelial cells were the most abundant cell types in saliva, consistent with previous literature and our reanalysis of recent single-cell RNA-seq data of human sputum tissue [48] (Figure S6). Comparing cell type proportions between diabetic and non-diabetic samples, we observed no significant changes in major cell types (Figure 4), except for a marginally significant difference in naïve T cells. However, this association was not significant after p-value adjustments. Our analysis also revealed that cell type proportions are highly correlated with the top Principal Components (PCs) of the DNA methylation matrix (Figure S7), emphasizing the dominant role of cell proportions in the epigenetic variability [33] and echoing the importance of including these variabilities in EWAS analysis to account for cell type heterogeneity [49].

**Figure 4:**
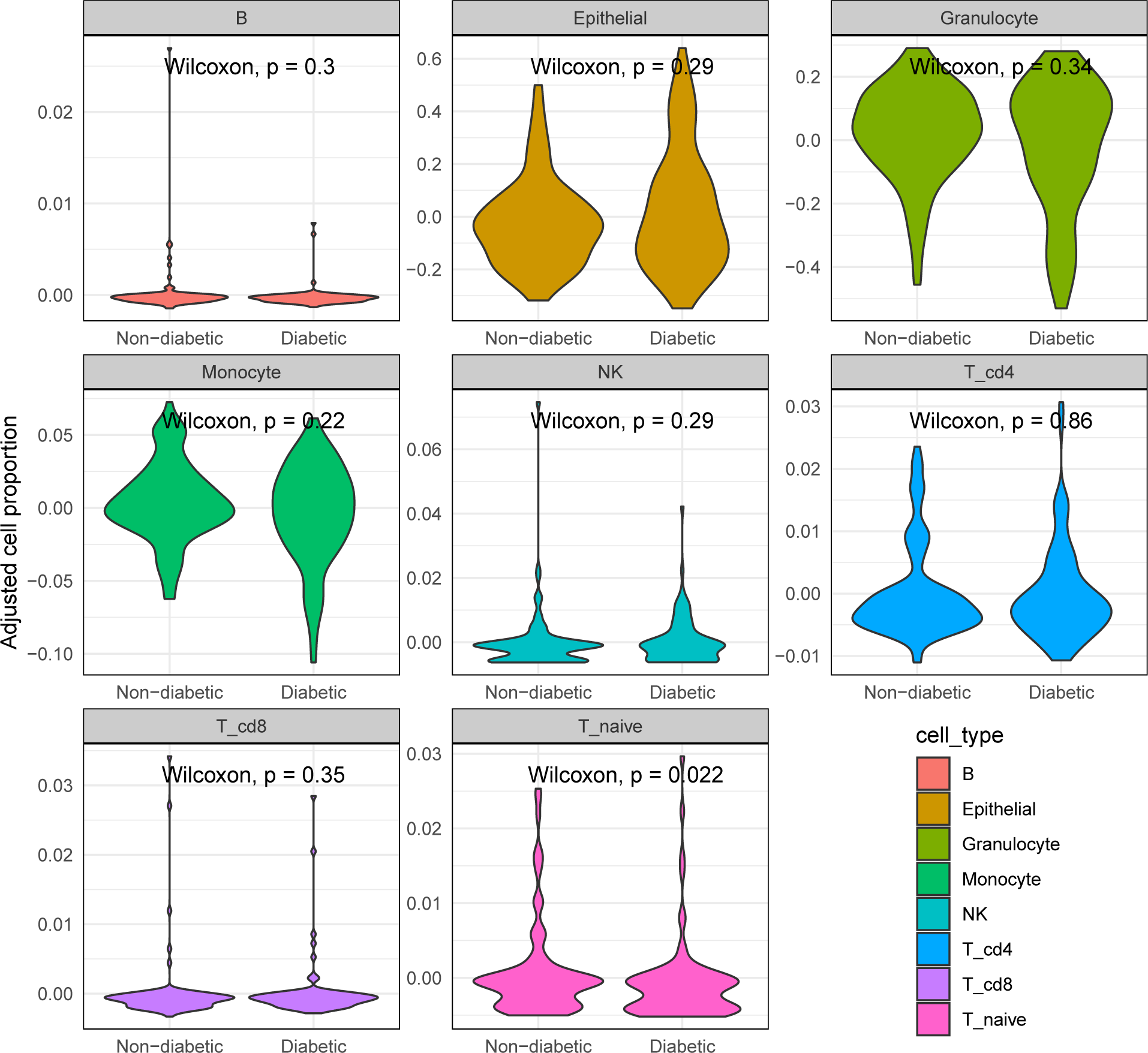
Differential cell type proportions between diabetic and non-diabetic samples. Violin plots show the difference in cell type proportions between diabetic and non-diabetic samples for each cell type after adjusting other covariates (age, sex, ethnicity, and batch). Wilcoxon p-values are annotated in each subplot, revealing no significant difference in cell proportions between the two groups, except a marginally significance for naïve T cell (p=0.022).

In conclusion, our analysis demonstrated that TBS sites capture cell-type information and enable accurate cell-type deconvolution. Notably, the similar cell type proportions observed between diabetic and non-diabetic groups suggest that diabetes-related epigenetic changes in saliva are driven by intrinsic molecular alterations rather than shifts in cell composition.

### EWAS reveals differential DNA methylation associated with T2D status

Another distinct advantage of TBS is its ability to elucidate the epigenetic mechanisms underlying diabetes at the molecular level, providing valuable insights into disease pathways and potential therapeutic targets. To demonstrate this potential, we conducted an epigenome-wide association study (EWAS) on the TBS data. In this analysis, we accounted for key covariates such as age, sex, ethnicity, study batches, and cell-type proportions, to mitigate the influence of confounding factors and identified CpG sites associated with diabetic states. The EWAS results, visualized with a Manhattan plot (Figure 5A) and a QQ plot (Figure S8), revealed 12 CpG sites significantly associated with T2D, with 7 of these sites near genes previously implicated in diabetes pathogenesis, such as *ABCG1* [32], *LDLRAD4* [50], and *TYK2* [51]. Figure 5B shows methylation level differences at the top CpG sites between diabetic and non-diabetic groups after adjusting for covariates.

**Figure 5:**
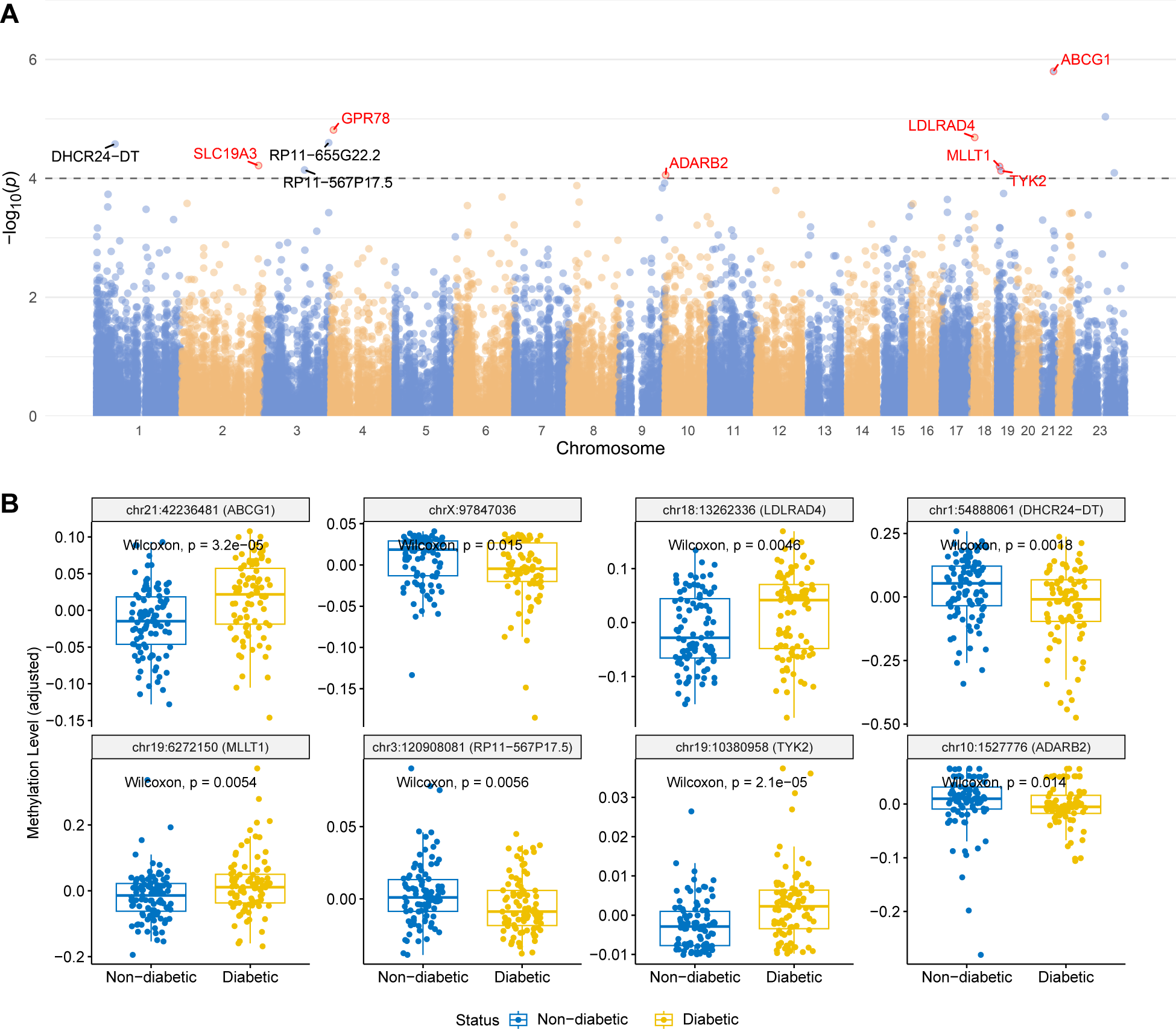
EWAS analysis identifies methylation sites associated with diabetes status. (A) Manhattan plot depicting the epigenome-wide association between DNA methylation levels and T2D status. Each dot represents a CpG site, with the −log10(p-value) plotted against its chromosomal position. The horizontal dashed line indicates the suggestive significance threshold (10*^−^*^4^). Genes located within a 2kb window of the top CpG sites are annotated, with established diabetes-related genes highlighted in red, such as *ABCG1*, *LDLRAD4*, *TYK2*, etc. (B) Boxplots illustrating the methylation levels (adjusted for covariates) at selected top CpG sites. Each plot panel compares the methylation levels between diabetic (yellow) and non-diabetic (blue) samples at the specific CpG site, highlighting their potential role in diabetes pathogenesis.

Notably, the strongest signal was observed in the *ABCG1* gene region, corroborating a recent meta-analysis of blood-based EWAS [32] that identified *ABCG1* as a top hit across five cohorts with over 3,000 samples. *ABCG1* plays a crucial role in regulating lipid metabolism and cholesterol efflux, which are essential for maintaining cellular lipid homeostasis [52]. The dysfunction of *ABCG1* is particularly detrimental in the context of diabetes, where impaired cholesterol efflux can exacerbate insulin resistance and promote atherosclerosis [53], a common complication of the disease. Furthermore, the accumulation of lipids can result in cellular stress and apoptosis [54], which in turn triggers an immune response and leads to chronic inflammation, further accelerating diabetes progression and increasing cardiovascular disease risk. The role of *ABCG1* in lipid regulation and its broader impact on inflammation and cell viability highlight its potential as a therapeutic target in diabetes management.

Our EWAS findings, particularly the significant signal at the *ABCG1* region, highlight the gene’s critical role in diabetes. These results validate the utility of saliva-based DNA methylation analysis in diabetes research and emphasize the potential of these epigenetic markers as biomarkers for diagnosing diabetes, predicting risk, and informing the development of targeted therapeutic strategies.

### Predictive performance of individual methylation sites for T2D status

To evaluate the potential of DNA methylation as a biomarker for diabetes diagnosis, we analyzed the predictive performance of individual methylation sites using ROC analysis. Figure 6 illustrates the ROC curves of all tested sites, with chr19:10380958 (*TYK2*) and chr21:42236481 (*ABCG1*) achieving AUC values of 0.683 and 0.681, respectively, indicating moderate predictive ability. The shaded region, representing the 95% quantile range of ROC curves across all sites, highlights the variability in predictive performance. These results demonstrate that while some individual sites show moderate performance, most exhibit weak signals, underscoring the importance of refining site selection. This validates the need for a targeted sequencing strategy, as it can effectively enrich informative loci, improving the signal-to-noise ratio and enabling precise and efficient methylation profiling.

**Figure 6:**
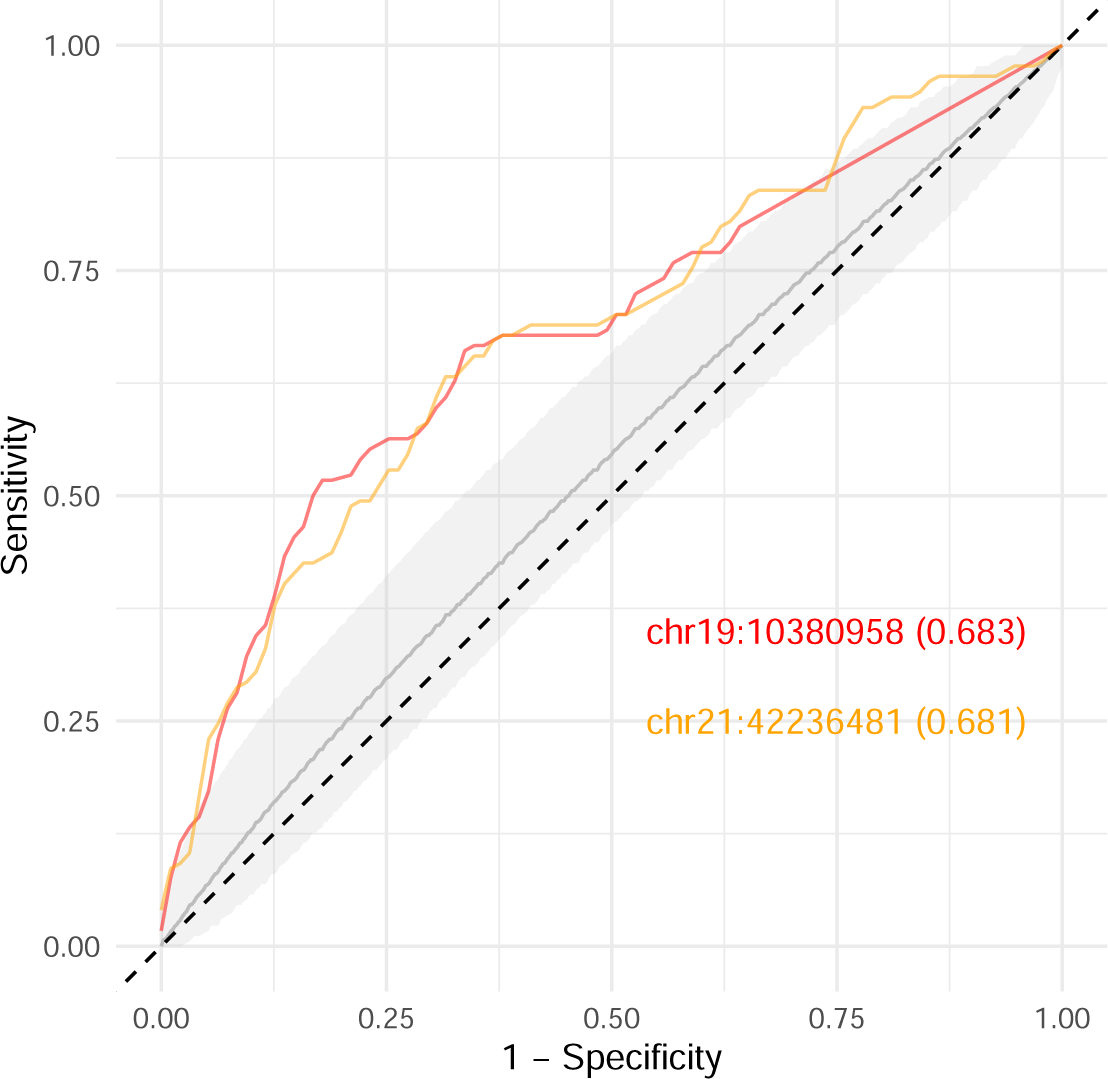
ROC curve for diabetes status classification using individual methylation sites. This ROC curve highlights the classification performance of two key methylation sites, chr19:10380958 and chr21:42236481, in predicting T2D status, with respective AUC values of 0.683 and 0.681. The shaded region denotes the 95% range of predictive performance across all other analyzed methylation sites, providing context for the highlighted sites’ relative performance, with the dashed diagonal line representing AUC 0.5 as a reference

Additionally, although the predictive power of individual loci is limited, combining methylation profiles within multivariate or ensemble frameworks offers a promising path forward. Future model development should focus on integrating information across multiple loci to enhance predictive accuracy and robustness. These strategies have the potential to yield reliable, clinically actionable tools for diabetes diagnosis and risk stratification, underscoring the transformative potential of the saliva DNA methylome as a scalable, non-invasive approach for advancing T2D biomarker discovery and improving disease management.

## Discussion

The rising prevalence of type 2 diabetes (T2D) underscores the need for innovative approaches that extend beyond traditional diagnostics to explore the molecular mechanisms underpinning the disease. Identifying reliable biomarkers and investigating epigenetic modifications, such as DNA methylation, can deepen our understanding of T2D pathogenesis, enable early detection, and inform the development of targeted therapeutic strategies. To address the need for accessible and noninvasive approaches, this study evaluated the potential of saliva DNA methylome for T2D biomarker discovery and diagnostic applications, offering insights into the molecular and cellular dynamics underlying the disease.

One key challenge in methylation profiling is the high cost of obtaining informative and accurate measurements. Whole-genome bisulfite sequencing (WGBS) provides comprehensive coverage but requires high sequencing depth, making it prohibitively expensive for large-scale studies. Methylation arrays, while more affordable, capture only a fixed, small subset of CpG sites, potentially overlooking critical variations relevant to disease. To overcome these limitations, we developed a cost-efficient two-step strategy, combining WGBS to identify key regions with targeted bisulfite sequencing (TBS) for high-depth profiling. This approach significantly reduces costs while maintaining precision, making it suitable for broader and cohort-level applications.

Using this combined strategy, we obtained compelling evidence supporting the use of saliva DNA methylation for T2D biomarker discovery and risk assessment. Through WGBS, we identified differentially methylated regions (DMRs) associated with T2D, particularly enriched in pathways related to immune response and metabolic regulation. These results align with existing blood-based studies [37], confirming that saliva, like blood, harbors diabetes-specific epigenetic signatures. The subsequent application of targeted bisulfite sequencing (TBS) enabled precise quantification of DNA methylation in these key regions at the cohort scale. Importantly, cell type deconvolution of the TBS data revealed no significant differences in cell proportions between diabetic and non-diabetic groups, suggesting that the observed methylation changes are primarily intrinsic rather than driven by shifts in cell composition. Further supporting these findings, an epigenome-wide association study (EWAS) conducted on the TBS data identified significant CpG sites, with the top hit in the *ABCG1* gene region, consistent with prior blood-based findings [32]. Collectively, our findings provide the first validation of T2D-specific methylation signals in saliva, establishing a novel paradigm for non-invasive diabetes screening and offering valuable insights into the epigenetic basis of this prevalent disease.

Despite these promising findings, our study has limitations that warrant further investigation. The relatively small sample size may have reduced the statistical power of our findings, potentially leading to missing important epigenetic signals. Expanding the sample size and including a more diverse population would enhance the robustness and generalizability of the results. Additionally, many diabetic participants were under good glycemic control, which may have attenuated the strength of detectable epigenetic changes. Future studies should include individuals at various stages of disease progression to capture a broader range of epigenetic variations. While our probe panel targeted diabetes-related sites, it could be further optimized by integrating prior knowledge to capture a wider range of diabetes-associated signals, particularly regions near genes involved in insulin signaling, glucose metabolism, and related pathways. Additionally, advanced machine learning approaches, such as ensemble and contrastive learning [55, 56], hold promise for enhancing diagnostic model performance by effectively integrating subtle signals linked to different disease states. Addressing these limitations through larger cohorts, refined probe designs, and advanced modeling techniques will be crucial for maximizing the potential of saliva DNA methylation in diabetes research and diagnostics.

Looking ahead, further research could greatly enhance the utility and impact of our approach. Advanced barcoding and multiplexing techniques, such as Time-Seq [57], could further reduce costs, making this method even more accessible for large-scale studies and routine clinical applications. The non-invasive nature of saliva collection, combined with cost-effective methylation profiling, offers a practical and scalable solution for diabetes screening and longitudinal monitoring. Conducting longitudinal studies will be critical to establish causal relationships between DNA methylation changes and T2D progression, providing deeper insights into disease mechanisms and enabling timely interventions. Ultimately, integrating saliva DNA methylation profiling into clinical practice has the potential to revolutionize diabetes diagnostics and monitoring, facilitating earlier detection, personalized treatment, and more effective disease management.

In conclusion, this proof-of-concept study validates diabetes-specific epigenetic signals in saliva, establishing saliva DNA methylation as a promising biomarker source for non-invasive T2D research and screening. By employing an innovative sequencing strategy that enhances precision while reducing costs, we have made epigenetic profiling feasible for large-scale studies and clinical applications. While further research with larger, more diverse cohorts is needed, this approach lays the groundwork for transforming diabetes diagnostics and monitoring, paving the way for more personalized and accessible care.

## Methods

### Sample collection and preparation

This study involved saliva samples collected as part of the Parkinson’s Environment and Genes (PEG) study [58, 59, 60]. While PEG is a case-control study focused on Parkinson’s disease (PD), the saliva samples utilized in this study were primarily unrelated to PD. Participants were recruited from various sources across three counties in the Central Valley of California (Kern, Fresno, and Tulare) during two study waves (2000-2007 and 2009-2015). Population controls were enrolled from the same regions using Medicare lists and residential tax assessor records. Demographic data, medical history, medication use, and lifestyle information were collected through standardized interviews. Saliva collection tubes were mailed to participants, who then returned them via shipping or during in-person examinations. For this study, samples from participants with and without type 2 diabetes were randomly selected from those available in the PEG study, ensuring that the diabetic and non-diabetic groups were matched for age, sex, and ethnicity (Supplementary Data 1). Two batches of 96-well plates were prepared: the first in 2020 (Diabetes n=48, Non-diabetic n=48) for Whole Genome Bisulfite Sequencing (WGBS), probe design, and a pilot Targeted Bisulfite Sequencing (TBS) study, and the second in 2022 (Diabetes n=42, Non-diabetic n=54) for an expanded TBS study. The batch was included as a covariate in the downstream analyses. Each individual’s saliva samples were sent to the UCLA Neuroscience Genomics Core (UNGC) for DNA extraction. Typically, 2.5 mL to 4 mL of saliva samples were collected using the Oragene saliva collection kit, followed by the standard manufacturer protocol of the Qiagen Puregene DNA extraction kit. After purification and extraction, the DNA concentration was measured using a NanoDrop 8000 spectrophotometer, and the extracted DNA samples were stored at −20°C before library preparation.

### Whole genome bisulfite sequencing (WGBS)

To optimize cost efficiency, the extracted saliva DNA samples from the first batch were aggregated into four groups, matched by age and sex, as detailed in Supplementary Data 1. Each grouped DNA was pooled and subjected to whole genome bisulfite sequencing (WGBS) following established protocols [61]. Specifically, one microgram of purified DNA was sonicated using the Bioruptor Pico (Diagenode) for 15 cycles of 30 seconds ON and 90 seconds OFF, targeting a fragment size of 200-300 bp. The NEB Next Ultra II DNA kit (New England Biolabs) was used for subsequent end-repair, A-tailing, and ligation of pre-methylated unique-dual indexed adapters (Integrated DNA Technologies, custom synthesis). Bisulfite conversion was performed with the EZ DNA Methylation-Gold kit (Zymo Research). Final library amplification (12 PCR cycles) was conducted using KAPA HiFi U+ polymerase (Roche Sequencing) and IDT xGen Primers. Library quality was assessed using the D1000 Assay on a 4200 Agilent TapeStation, and concentrations were quantified with the Qubit dsDNA BR Assay (Life Technologies). Sequencing was conducted on a NovaSeq 6000 platform (S4 lane), generating paired end reads of 150 base pairs.

### WGBS data processing and DMR analysis

The raw sequencing reads underwent quality control using FastQC [62], followed by adapter and low-quality base trimming with fastp [63]. The trimmed reads were aligned to the reference genome (hg38) using BSBolt [64], with PCR duplicates marked with samtools [65]. Methylation levels of CpG sites were quantified for each sample, then aggregated into a methylation matrix. For downstream analysis, only sites with at least five counts in all four pooled samples were retained. The methylation matrix is available in Supplementary Data 2.

Differentially methylated region (DMR) analysis of the WGBS data was conducted using metilene [66] (version 0.2-8), with each candidate region required to contain a minimum of five CpG sites. In total, 162,833 genomic regions were analyzed. The statistical significance of methylation differences between diabetic and non-diabetic groups was evaluated using the Mann-Whitney U-test and the 2D Kolmogorov-Smirnov test for each region. Regions were considered significantly differentially methylated if they exhibited p-values below 0.01 for both tests and an absolute methylation difference exceeding 0.2 between the two groups. Supplementary Data 3 provides a comprehensive list of all candidate regions and identified DMRs, including their genomic coordinates, absolute methylation differences, and statistical significance levels.

### Genomic region enrichment analysis and probe design

Following the identification of differentially methylated regions (DMRs) between diabetes and non-diabetes WGBS data, we conducted a genomic region enrichment analysis using the R package rGREAT [67] (version 2.4.0) in online mode. This analysis compared the DMRs against the total examined genomic regions as background, revealing significant enrichment patterns and biological relevance of the observed methylation changes. The identified DMRs were later submitted to Integrated DNA Technologies (IDT, https://www.idtdna.com/) for probe design, resulting in 937 custom probes targeting these regions. To further understand the regulatory context, we performed motif enrichment analysis using HOMER [68] (version 4.11), identifying enriched transcription factor binding sites (TFBS) for the probe-enriched regions.

In addition to the newly designed probes, we also incorporated previously designed probes targeting regions of interest from earlier studies [42, 43]. These probes were selected based on loci identified in public epigenome-wide association studies (EWAS) related to aging, cell types, and metabolic disorders. Due to an update in the probe set, there are slight differences between the probes used in batch 1 and batch 2, each containing a small set of extended probes labeled as ‘batch1 extended’ and ‘batch2 extended.’ The probes consistently used throughout the TBS study are collectively labeled as the ‘Total Panel’ and referred to as ‘total probes’ throughout the manuscript. The complete panel of probes, including both the Total Panel and extended probes, is detailed in Supplemental Data 4, with their sequences and target regions provided.

### Targeted bisulfite sequencing (TBS)

For targeted bisulfite sequencing (TBS), 250 to 500 ng of purified gDNA from each sample was fragmented, and libraries were constructed following the same procedure as described in the WGBS protocol. Groups of 16 libraries, each with a unique dual index adapter, were pooled together, concentrated via SpeedVac, and subjected to targeted enrichment using custom 5’-biotinylated probes (IDT, xGen Custom Hybridization probe panel) (Supplemental Data 4). Enrichments were performed with the xGen Hybridization Capture kit (IDT), following the manufacturer’s instructions, including overnight hybridization at 65°C. Bisulfite conversion of captured DNA was conducted using the EZ Methylation Gold kit (Zymo Research). Final PCR amplification employed KAPA HiFi Uracil+ (Roche) with the following conditions: initial denaturation at 98°C for 2 minutes, followed by 16 cycles of 98°C for 20 seconds, 60°C for 30 seconds, and 72°C for 30 seconds, with a final extension at 72°C for 5 minutes. PCR products were purified using SPRI beads, and library quality control was conducted with the High-Sensitivity D1000 Assay on the 4200 Agilent TapeStation. Pools of 96 libraries were sequenced on a NovaSeq 6000 with paired-end 150-base reads.

### TBS data processing and quality control

The raw sequencing reads of TBS data underwent a standardized preprocessing pipeline, including quality control, trimming, alignment, PCR duplicate marking, and methylation calling, as outlined in the WGBS data processing protocol. The methylation level of each CpG site is computed as follows:

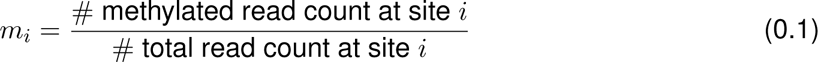

To ensure data quality, samples with fewer than 2.5 million unique reads post-PCR deduplication or identified as outliers through PCA on the methylation level matrix were excluded from further analysis. After the data quality control, a total of 182 samples (Diabetic n=87, Non-diabetic n=95) were retained for further investigation.

We also performed quality control on the features. For epigenome-wide association studies, we focused on count data, retaining only those sites with read counts exceeding 10 in at least 80% of the samples. For analyses focused on methylation levels, such as cell deconvolution or machine learning model development, we retained only those sites that had at least 20 counts in at least 80% of the samples to ensure a reliable methylation level estimate. Missing values in the methylation level matrix were imputed using the KNN algorithm (k=5) implemented by R package impute [69] (version 1.70.0).

### Cell type deconvolution

To ensure deconvolution accuracy, we first analyzed the cell composition in saliva using a single-cell RNA-seq dataset (GSE158055) [48] from the CELLxGENE Discover Data Portal (https://cellxgene.cziscience.com), confirmed the predominance of epithelial and immune cells (Figure S6A-B). Building on this confirmation, we then compiled a comprehensive cell type methylation reference for deconvolution by integrating Whole Genome Bisulfite Sequencing (WGBS) profiles from the DNA methylation atlas (GSE186458) [46]. This reference encompasses epithelial and key immune cell types—granulocytes, monocytes, NK cells, B cells, and T cells (CD4, CD8, and naïve)—with detailed accession IDs and labels provided in Supplementary Data 5. Cell type-specific differentially methylated regions (DMRs) were identified by comparing each cell type against all others using metilene [66] (version 0.2-8), with parameters aligned to prior DMR analyses. Regions with a methylation difference exceeding 0.3 and an adjusted p-value below 0.05 (Benjamini-Hochberg correction) were extracted, and CpG sites in these regions were used to create the cell type methylation signature matrix. The resulting matrix, which serves as a robust reference for deconvolution, is available in Supplementary Data 6.

Given the limited number of CpG sites captured by TBS data, we further validated the deconvolution accuracy on TBS data using synthetic DNA methylation profiles. We generated 100 in-silico samples by mixing DNA methylation profiles of cell types with known proportions, with random gaussian noise (mean = 0, standard deviation = 0.05) added to mimic the variability in methylation levels due to sequencing noise. These synthetic datasets were then filtered to include only the CpG sites presented in the TBS methylation level matrix. Deconvolution was performed using the Houseman method [47], a well-established NNLS (non-negative least squares) approach for estimating cell-type compositions from bulk methylation data. The deconvolution accuracy was assessed by comparing the estimated and true cell proportions using key metrics, including R-squared and root mean square error (RMSE), confirming the precision and reliability of the method for TBS sites. This validated framework was then applied to deconvolve cell-type compositions in bulk saliva samples. Detailed cell proportions for each sample are provided in Supplementary Data 7.

### Epigenome-wide association study

To prioritize the risk CpG sites associated with T2D, we conducted an epigenome-wide association study (EWAS) using the methylation read counts data using R package DSS [70]. Specifically, the DSS package utilized a beta-binomial modeling strategy to model the methylated counts *Y_i_* based on the total counts *N_i_* for site *i* by

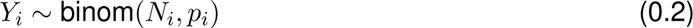

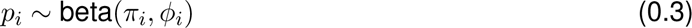

where *π_i_*, *ϕ_i_* are the mean and dispersion parameter for site *i*. The mean parameter was modeled as 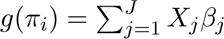 where *g*(·) is the link function and *X_j_* for *j* = 1*,…, J* are the covariates (including the variable of interest and other covariates). By testing the coefficient *β_j_* = 0 using the F-test, the significance level of association between diabetes status and methylation of site *i* can be assessed. For our analysis, we used the methylation count matrix for the EWAS analysis, including age, sex, ethnicity, batch, and cell proportions as covariates, and tested whether a site is associated with diabetes. A Manhattan plot is used to visualize the testing results. Genes within 2kb window of the most prominent sites that passed the with suggestive p-value (10^−4^) is annotated on to the plot. Detailed methylation count matrix and test result are available in Supplementary Data 2.

**Table 1:**
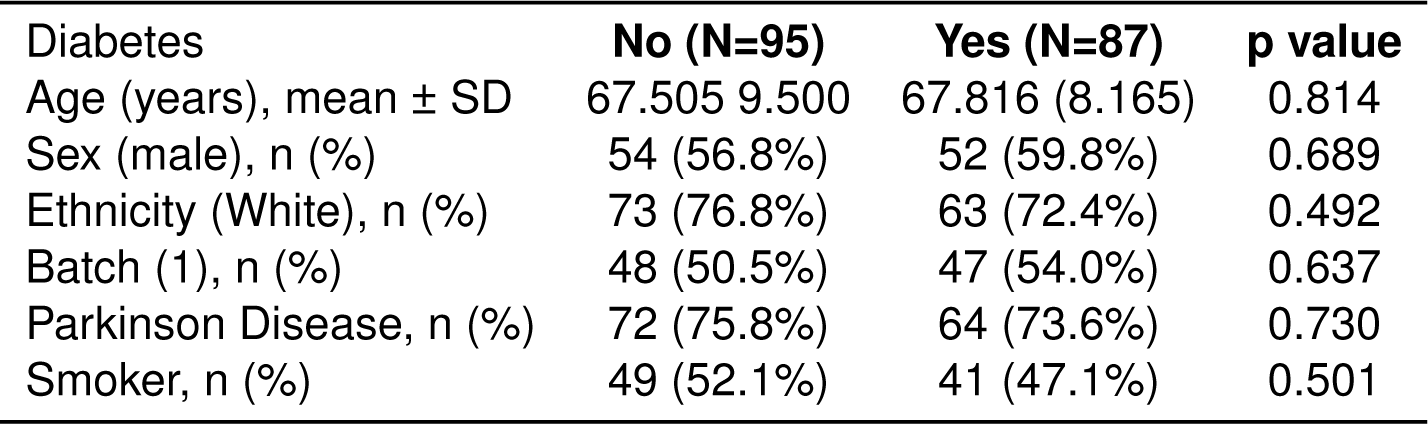
Characteristics of the study population.

## Data availability

The raw data will be released upon manuscript publication.

## Acknowledgements

We would like to thank all the participants who donated samples and make this study possible. We appreciate Dr. David Wong from School of dentistry at UCLA for his valuable insights and discussion. We also thank the UCLA Neuroscience Genomics Core (UNGC) and Broad Stem Cell Sequencing Core (BRSRC) for their help in DNA extraction, library preparation, WGBS and TBS data generation. Figure 1 was created with BioRender.com, thus acknowledging it by courtesy.

## Author contributions

W.G., M.M., and M.P. conceived the research idea. W.G. performed downstream data analysis and wrote the manuscript with the help of M.M. and K.C.P. M.M. conducted the WGBS experiment, initiated the WGBS analysis and probe design, and edited the manuscript. M.T. helped with the data preprocessing and manuscript editing. K.C.P. and B.R. contributed to sample preparation, data collection, and manuscript editing. M.P. supervised the entire research.

## Supplementary Figures

**Figure S1:**
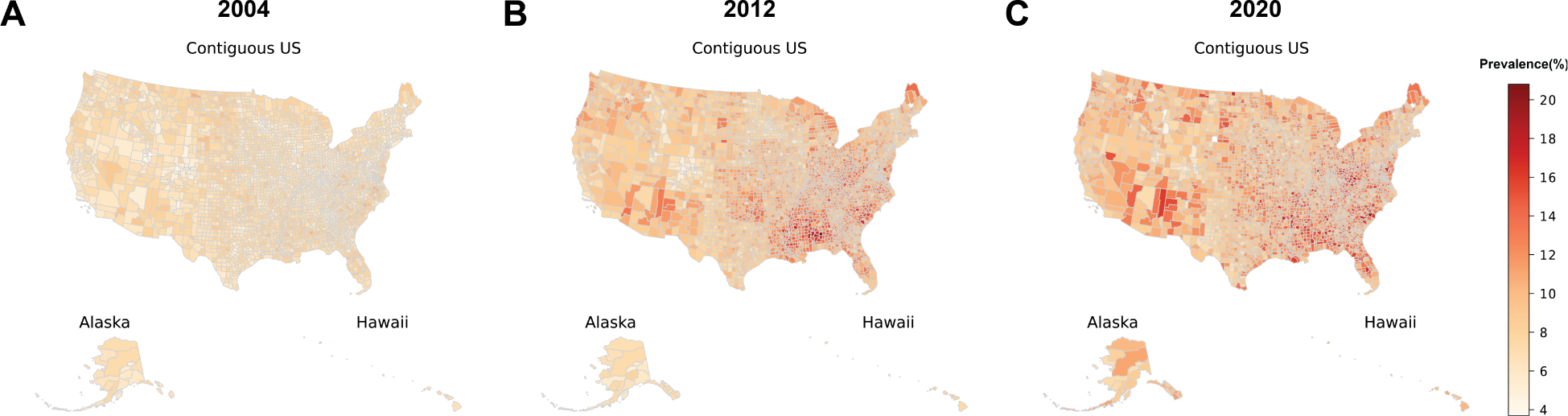
The increasing prevalence of diabetes across U.S. counties from 2004 to 2020. Choropleth map displaying the escalating diabetes prevalence in U.S. counties from 2004 (A), through 2012 (B), to 2020 (C), which underscores the growing public health challenge and the need for targeted interventions. The color gradient indicates the percentage of the population with diabetes, with darker colors representing increasing prevalence, as shown in the accompanying legend (4% to 20%). County-level diabetes prevalence data was obtained from the United States Diabetes Surveillance System (https://gis.cdc.gov/grasp/diabetes/DiabetesAtlas.html).

**Figure S2:**
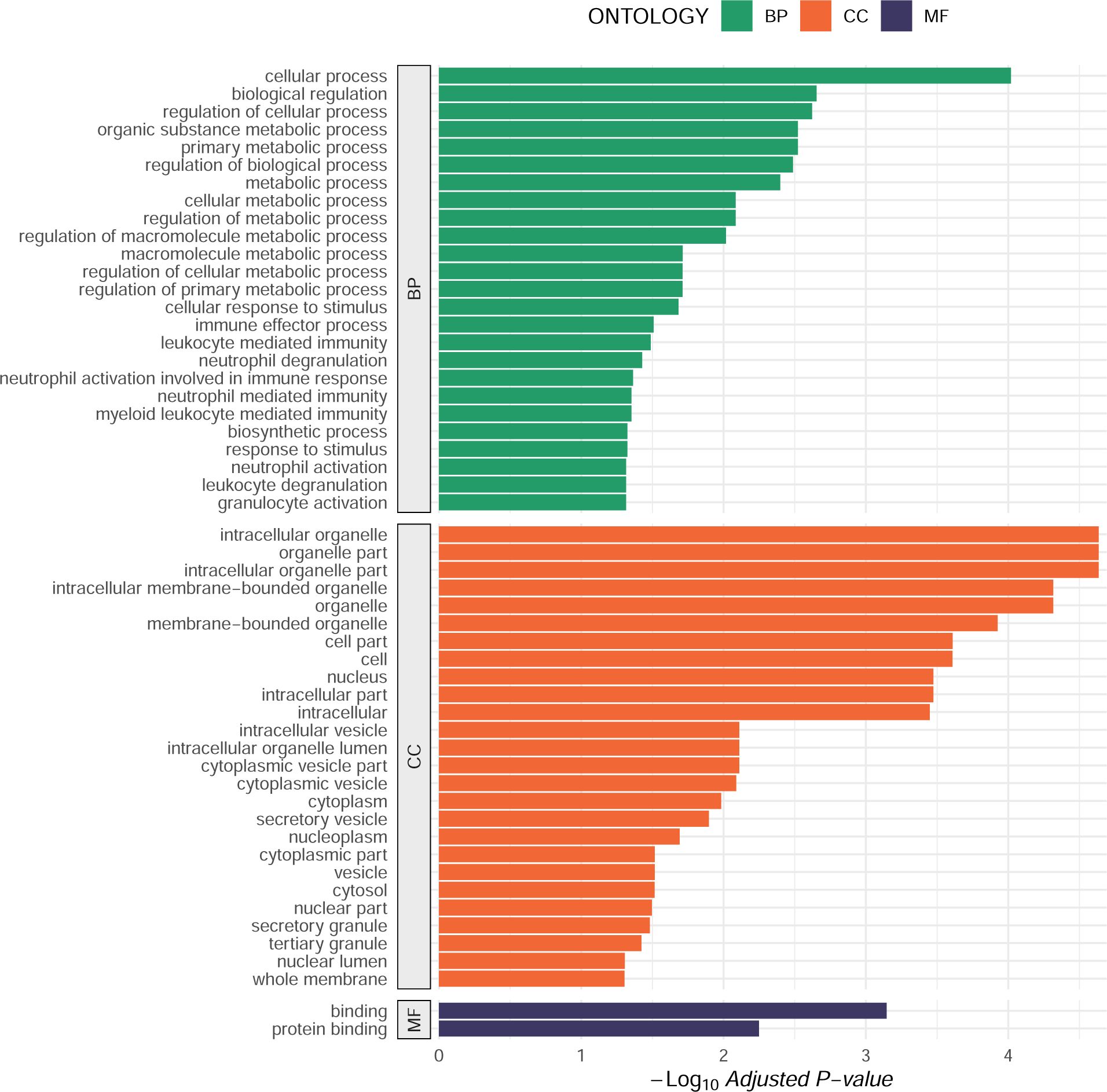
GO pathway enrichment for DMR regions in WGBS analysis. Genomic region enrichment analysis for differentially methylated regions (DMRs) identified in the Whole Genome Bisulfite Sequencing (WGBS) data. The bar plot presents enriched GO terms categorized by Biological Process (BP), Cellular Component (CC), and Molecular Function (MF) ontologies, with the x-axis showing the −log10 of the adjusted p-values. Only GO terms with an adjusted p-value below 0.05 are displayed. The analysis highlights significant associations of DMRs with various biological processes, particularly those related to metabolic functions and immune responses.

**Figure S3:**
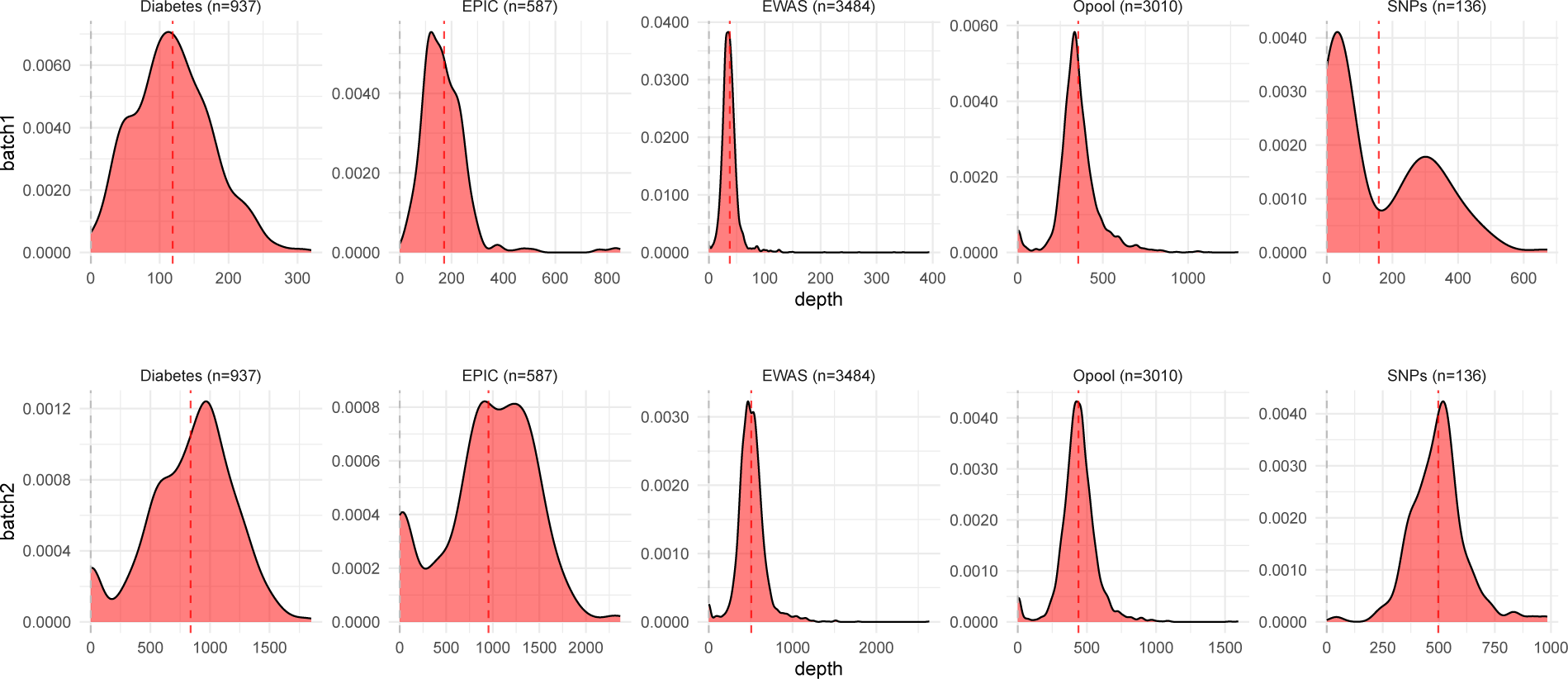
Depth distribution of targeted regions across probe sets in TBS. Density plots illustrate the depth distributions of different probe sets across two sample batches (upper panel: batch1, lower panel, batch2). Each subplot corresponds to a specific probe group—Diabetes, EPIC, EWAS, Opool, and SNPs—with the number of probes indicated in parentheses. Red dashed lines indicate the average depth for each probe set, with grey lines showing non-enriched background regions, underscoring the high efficiency of target enrichment achieved by targeted bisulfite sequencing (TBS).

**Figure S4:**
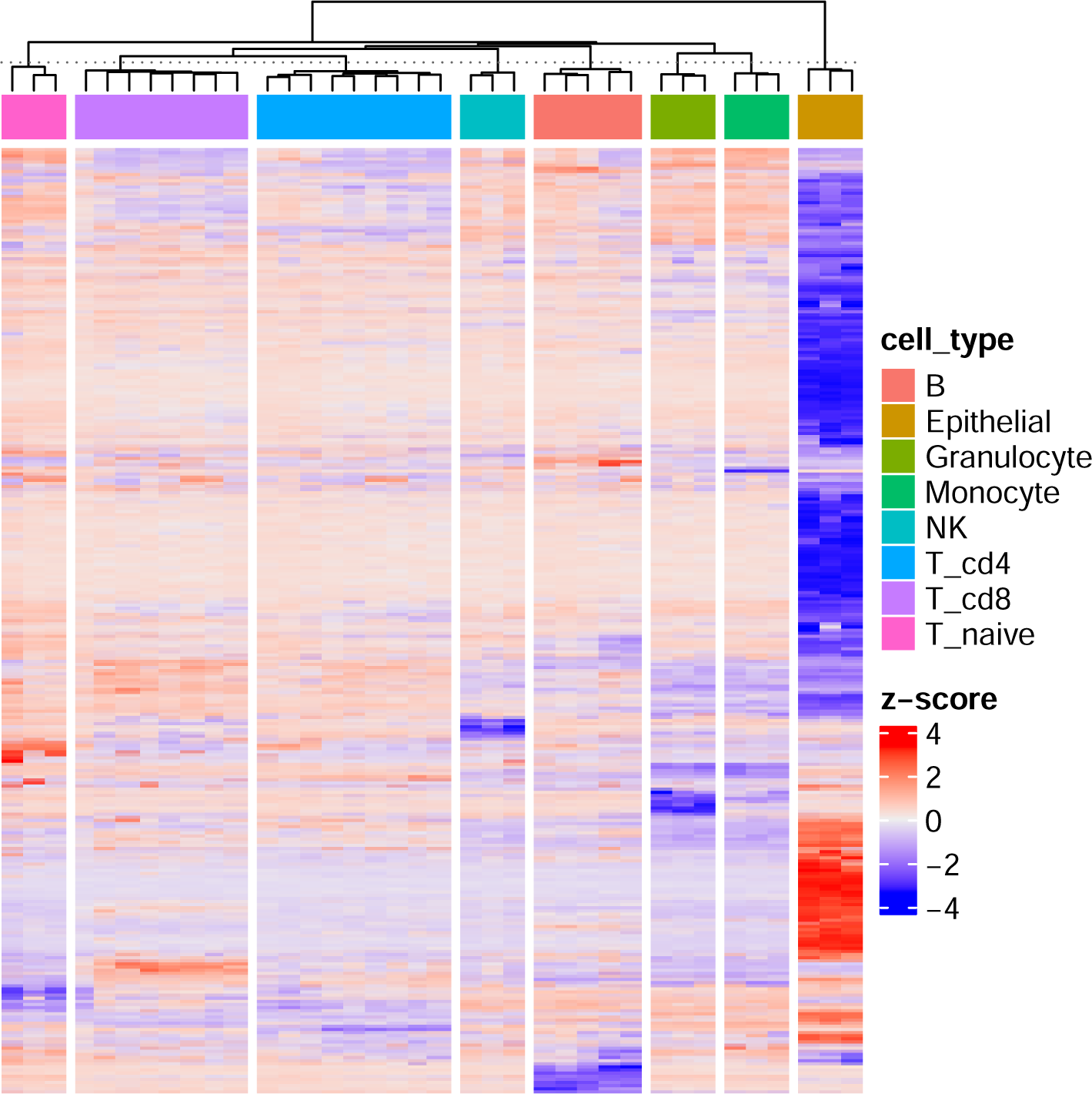
Cell-type specific methylation signatures at TBS sites. Heatmap showing the methylation profiles of various cell types, restricted to CpG sites that overlap with targeted bisulfite sequencing (TBS) data. Each column represents a sample from a specific cell type, with cell types indicated by the color bar at the top: B cells, epithelial cells, granulocytes, monocytes, NK cells, cd4+, cd8+, and naive T cells. The z-scores reflect relative methylation levels, with red indicating hypermethylation and blue indicating hypomethylation. The distinct clustering patterns in the heatmap confirm that TBS sites retain sufficient cell type identity information, allowing for a clear distinction between cell types. This demonstrates the efficacy of TBS in capturing cell type-specific epigenetic signatures, reinforcing its utility for studying cellular heterogeneity.

**Figure S5:**
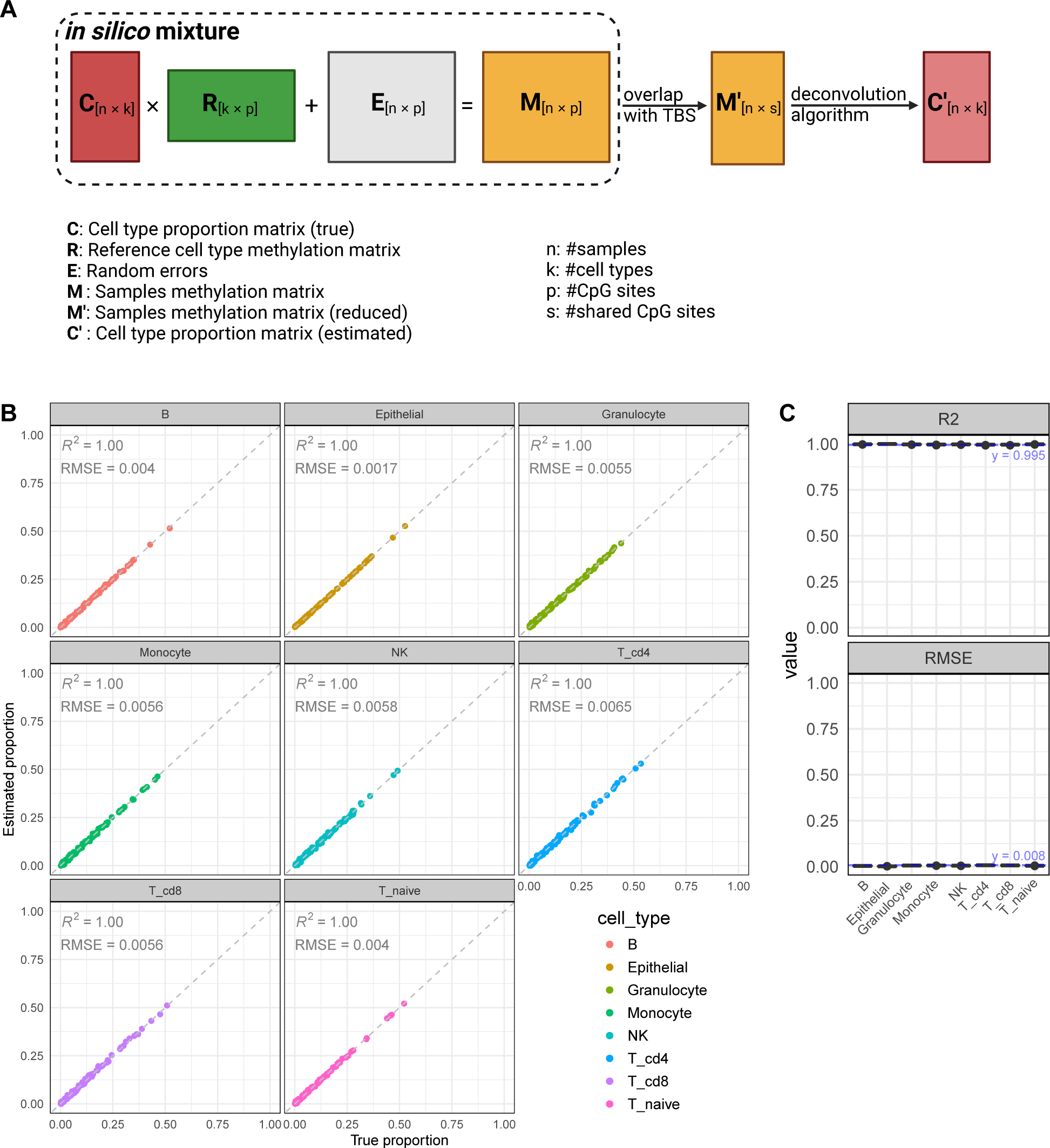
Simulated validation of TBS sites for accurate cell deconvolution. Simulation of cell deconvolution using targeted bisulfite sequencing (TBS) sites to confirm their support for accurate cell type estimation. (A) Schematic of the simulation workflow for cell deconvolution. The process begins with a true cell type proportion matrix, C, and a reference cell type methylation matrix, R, which are combined with random errors, E, to generate a simulated methylation matrix, M. This matrix is then refined to include only the sites overlapping with TBS data, creating a reduced methylation matrix, M’. A deconvolution algorithm is subsequently applied to estimate cell type proportions, C’, from the reduced methylation matrix. The accuracy of the deconvolution is then evaluated by comparing these estimated proportions with the true proportions. (B) Scatter plots showing the deconvolution accuracy across different cell types in a single simulated exaperiment, demonstrating high deconvolution accuracy. (C) Results from repeating the simulation 100 times, consistently showing high *R*^2^ values and low RMSE across all cell types, confirming that TBS sites robustly support accurate cell type deconvolution. (*R*^2^: coefficient of determination; RMSE: Root Mean Squared Error)

**Figure S6:**
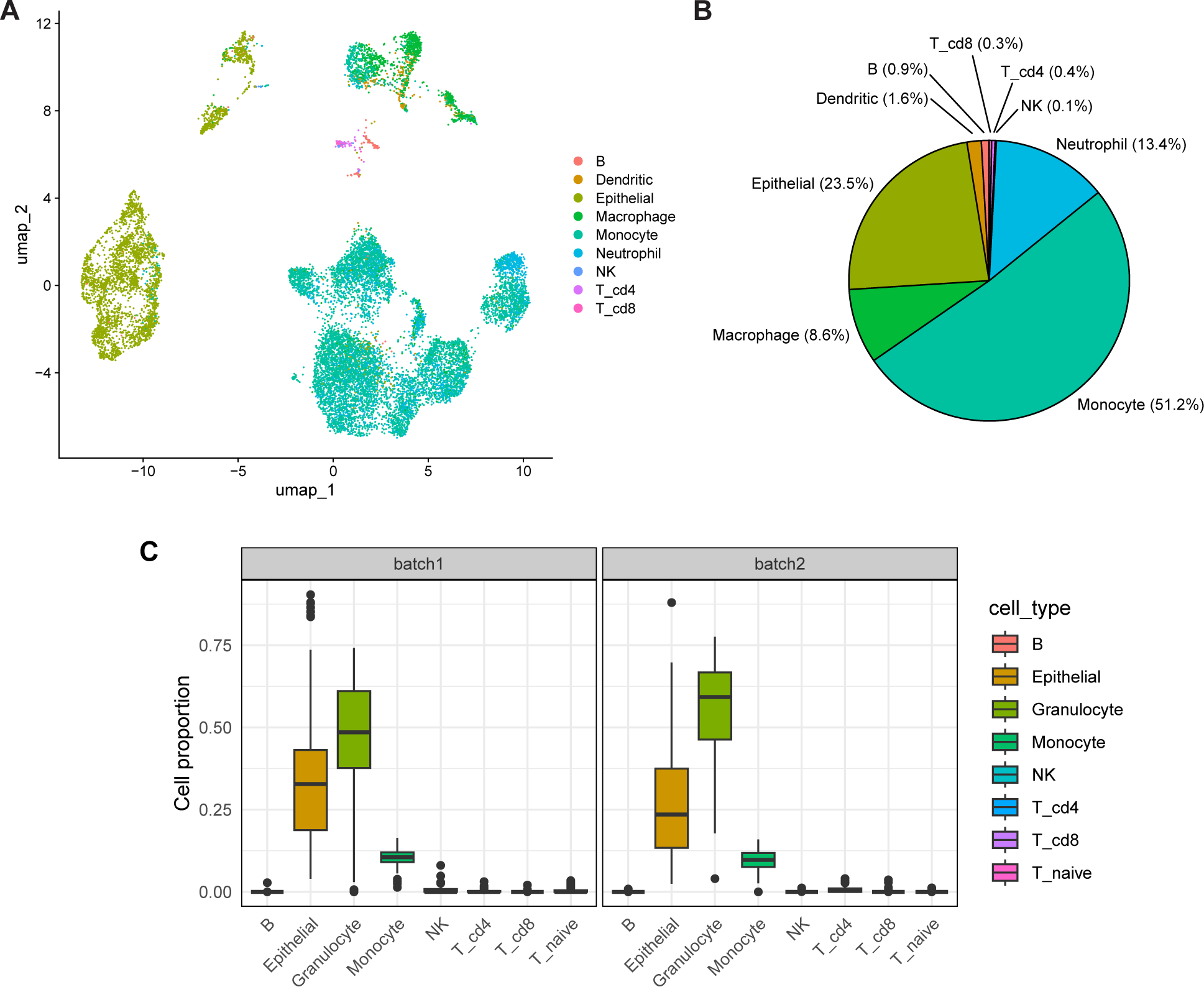
Cell type compositions in saliva: reanalysis of existing scRNA-seq dataset and TBS deconvolution results. (A-B) Cell type composition in saliva was revealed by the reanalysis of a previous single-cell RNA sequencing dataset from human sputum [48]. (A) UMAP plot displaying distinct clusters of cells, each colored according to its identified cell type. (B) Pie chart showing the abundance of cell type proportions, with Monocytes, Epithelial cells, and Neutrophils being the most abundant, followed by smaller populations of other immune cells. The reanalysis results validate the major cell types, confirming that immune cells and Epithelial cells are predominant in saliva samples. (C) Boxplots illustrating the cell type proportions in two batches (batch 1 and batch 2) derived from deconvolution analysis of bulk TBS data, highlighting the reproducibility across different batches. The deconvolution results show a similar pattern to the scRNA-seq findings, with Granulocytes, Monocytes, and Epithelial cells constituting most of the cell population. In contrast, other immune cells are present in lower proportions. The alignment between the scRNA-seq reanalysis and TBS data deconvolution results supports the reliability of deconvolution analysis. The differences in quantitative proportions may be attributed to inherent sample variation and technology biases, such as the scRNA-seq conducted on sputum from COVID-19 patients, which could have altered cell proportions and capture preferences.

**Figure S7:**
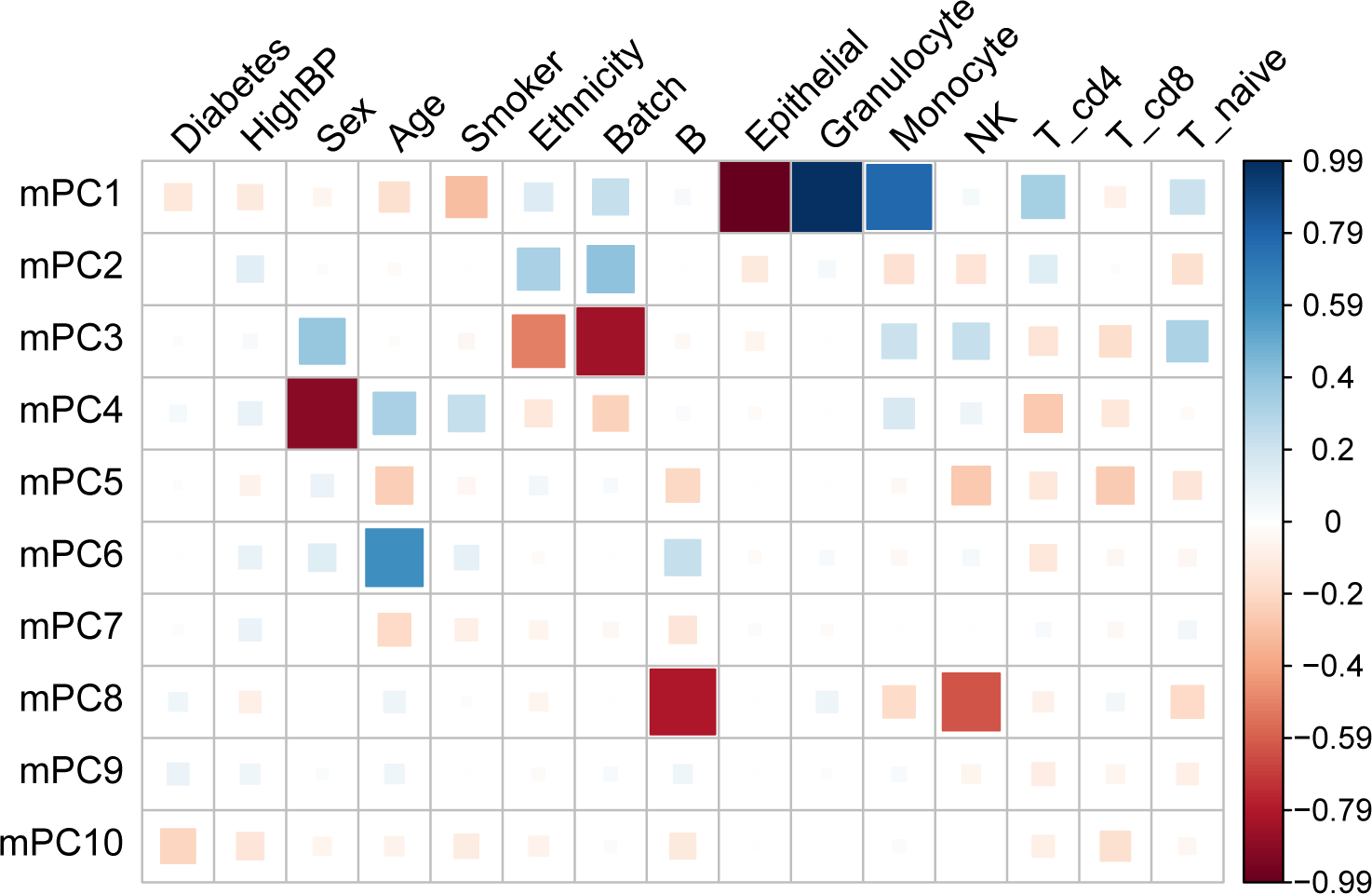
Correlation heatmap between methylation principal components (mPCs) and demographical and cellular variables. Heatmap illustrating the correlations between the top 10 methylation principal components (mPC1 to mPC10) and various demographical and cellular proportions. The color intensity and size of the squares represent the strength of the correlation, with blue indicating positive correlations and red indicating negative correlations, as shown by the color scale on the right. These correlations suggest that sex, age, ethnicity, and cell proportions are dominant factors of DNA methylation variations in the TBS data.

**Figure S8:**
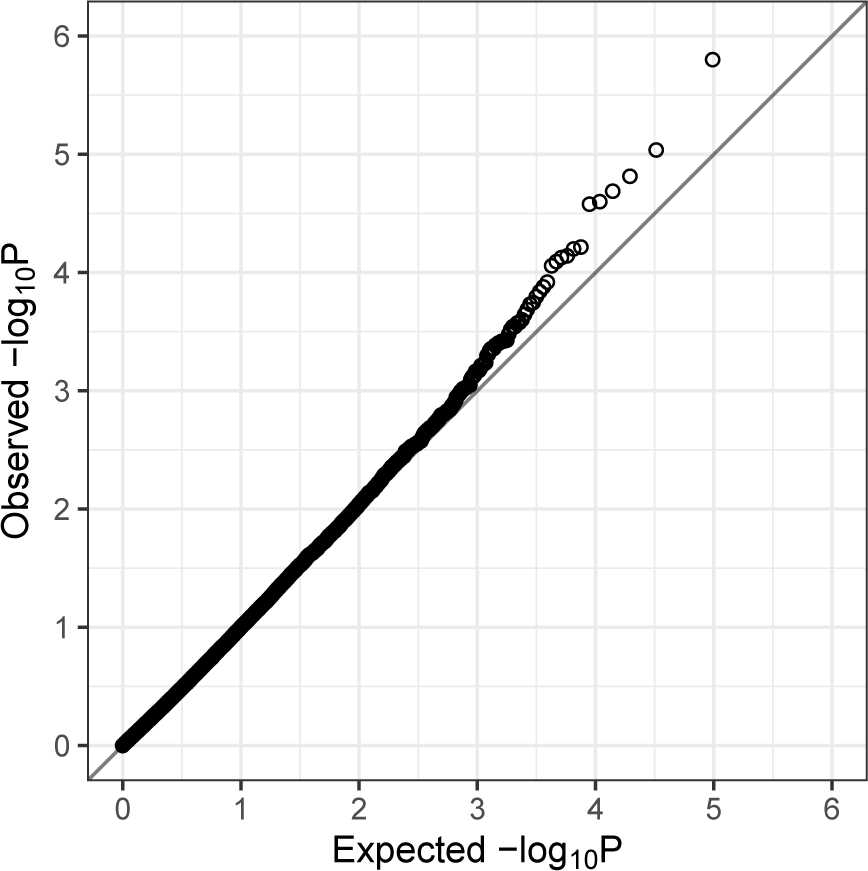
Quantile-Quantile (Q-Q) plot for EWAS analysis. The Q-Q plot compares observed −log10(p-values) from the EWAS with expected values under the null hypothesis. Points along the diagonal indicate concordance between observed and expected p-values, while deviations from the diagonal, particularly at the upper tail, suggest the presence of CpG sites with significant associations that exceed what would be expected by chance. The plot shows a slight deviation from the diagonal in the higher −log10(p-value) range, indicating the presence of true associations in the dataset.

